# Continuum of Core 1 Biomarkers in Preclinical Alzheimer’s Disease

**DOI:** 10.1101/2025.09.17.25336007

**Authors:** Leonardino A. Digma, Christina B. Young, Joseph R. Winer, Karly A. Cody, Kyan Younes, Jintao Sheng, Philip S. Insel, Robert A. Rissman, Reisa Sperling, Elizabeth C. Mormino

## Abstract

**Background and Objectives:** Biological Staging for Alzheimer’s disease (AD) in clinically unimpaired (CU) individuals is critical for early detection efforts. In this study, we evaluated whether Core 1 biomarkers (plasma ptau-217 and amyloid-PET) within Biological Stage A, the earliest biological stage of AD, predicts progression of downstream biomarkers and cognition.

**Methods:** We used baseline plasma ptau-217 and amyloid-PET, and longitudinal tau-PET, atrophy, and cognition data from the recently completed Anti-Amyloid Treatment in Asymptomatic Alzheimer’s (A4) Study. PET data were used to identify participants within AD Biological Stage A (amyloid-PET positive and medial temporal tau-PET negative). Within these Stage A participants, linear mixed effects models were used to examine associations between continuous baseline levels of plasma p-tau217 and amyloid-PET burden with longitudinal regional tau-PET, atrophy, and cognition. We additionally evaluated whether continuous p-tau217 and amyloid-PET burden within this group was associated with higher risk of progression to Biological Stage B+ (tau-PET positive in the medial temporal lobe). In our statistical models, we included covariates for age, sex, and APOE4 carriage.

**Results:** Of 335 A4 participants with complete biomarker data, 222 were identified as being in Biological Stage A. Among Biological Stage A CU, continuous baseline plasma p-tau217 and amyloid-PET burden were independently associated with faster tau-PET accumulation and atrophy in AD-relevant regions (mean follow-up time for both tau-PET and MRI: 4.2 years), as well as faster cognitive decline (mean follow-up time for PACC: 5.7 years) (all p<0.05). Plasma p-tau217 and amyloid-PET burden were independently associated with higher risk of progression to Biological Stage B+.

**Discussion:** In CU individuals, early changing AD biomarkers during the initial stage of AD (Biological Stage A) provide prognostic information of downstream markers of disease. Evaluation of the utility of these continuous measures in a real-world setting is warranted.

**Clinical Trial Information:** The A4 study was submitted for registration to clinicaltrials.gov on December 6th, 2023. The study is registered with ID NCT02008357. Screening and data collection for the study began in April 2014.

## Introduction

Alzheimer’s disease (AD) is defined by the presence of amyloid plaques and tau tangles in the brain^1^. Advances in biomarker technologies have enabled the detection of these protein aggregations during life with measures from cerebrospinal fluid (CSF), positron emission tomography (PET) brain imaging, and, more recently, plasma. The ability to measure the pathological hallmarks of disease in vivo has motivated multiple biological frameworks and staging schemes for AD. An updated framework was recently published by a workgroup of stakeholders^2^, proposing a sequence of Biological Stages (A-D) along the AD continuum based on biomarker profiles.

The 2024 biological criteria does not provide a specific guidance on the operationalization of the proposed biological framework. Rather, it proposes a flexible schema that can be applied based on available biomarker information. Broadly, it suggests a set of early changing “Core 1” biomarkers that, if positive, places a patient on the AD continuum (i.e., at least Biological stage A). Biomarkers deemed appropriate for Core 1 include measures of amyloid (A, amyloid-PET, CSF or plasma measures of Aβ 42) and early changing measures of tau (T_1_, CSF or plasma measures of p-tau181, 217, or 231). Individuals with a positive Core 1 biomarker can be further staged with “Core 2” biomarkers of disease prognostication, such as tau-PET (T_2_). Elevated tau-PET signal in the medial temporal lobe (MTL) is proposed to define Stage B, whereas moderate and high levels of neocortical tau-PET are recommended for Stage C and Stage D, respectively. Currently, tau-PET is the only recommended Core 2 biomarker that has sufficient data supporting its utility in the biological framework as a prognostication marker. Newer T_2_ biomarkers, such as MTBR-tau243^3^, are noted for their initial promise but warrant further validation.

As noted above, the earliest biological stage in the AD continuum is Biological Stage A and an individual can meet criteria for this stage by having an abnormal Core 1 biomarker (e.g., elevated amyloid-PET or plasma p-tau). An open question, however, is whether these early changing Core 1 biomarkers provide additional information beyond just binary statuses during the earliest stages of disease. In other words, when restricted to amyloid+ yet tau-PET negative participants (Biological Stage A), do continuous levels of Core 1 biomarkers predict subsequent changes relevant for disease progression. While biomarker measures of amyloid tend to show a bimodal distribution, there is evidence that meaningful variability exists beyond binary positive and negative status in cognitively unimpaired (CU) individuals^4^. Although plasma ptau-217 is a newer marker, there is also data demonstrating that variability in this measure can reveal groups at higher risk for disease progression ^5^. However, studies examining the continuum of early changing Core 1 biomarkers^5–7^ do not exclude tau-PET+ individuals, and it is possible that reported effects of plasma p-tau217 and/or continuous amyloid-PET burden are driven by the inclusion of this subgroup that is further along the AD trajectory (i.e., Stage B or higher).

We therefore sought to determine whether continuous values across two established Core 1 biomarkers—amyloid-PET and plasma p-tau217—were associated with subsequent disease relevant changes among Biological Stage A, CU individuals. Although these markers are classified as Core 1 in the new framework and are shown to correlate strongly across the AD spectrum^8–10^, they nevertheless measure distinct aspects of disease biology and may provide unique information during the earliest stage of AD. To evaluate the prognostic value of these early markers we applied the new staging criteria in a large sample of CU older adults, and determined whether the continuum of Core 1 biomarkers was predictive of subsequent longitudinal change in downstream disease markers within Biological Stage A.

## Methods

### Standard Protocol Approvals, Registrations, and Patient Consents

The A4 study was carried out in concordance with the guidelines from the International Council for Harmonisation Good Clinical Practice. The local ethics review board from each study site approved the study protocol. The study protocol is available as supplementary material in the primary publication for the A4 study^11^. Screening and data collection for the study began in April 2014. The study was completed in June 2023. All study participants provided written informed consent.

### Study Participants

Participant demographic, biomarker, and neuropsychological data were downloaded on November 5th, 2024 (a4studydata.org). We used data from participants that were enrolled in the A4 clinical trial (variable *SUBSTUDY*, value ‘A4’) and had amyloid-PET, plasma p-tau217, tau-PET, and MRI data available (Figure 1). The inclusion and exclusion criteria for the A4 trial have been described in detail previously^11^. Briefly, patients with normal cognition and evidence of amyloid in the brain, by a combination of visual and quantitative measurements, were enrolled in A4 and then randomized to either solanezumab or placebo. A subset of participants that were not eligible for A4 due to not having elevated brain amyloid were enrolled in the LEARN study. Data from participants enrolled in the LEARN study (variable *SUBSTUDY*, value ‘LEARN’) that had tau-PET data available were included (Figure 1).

**Figure 1.**
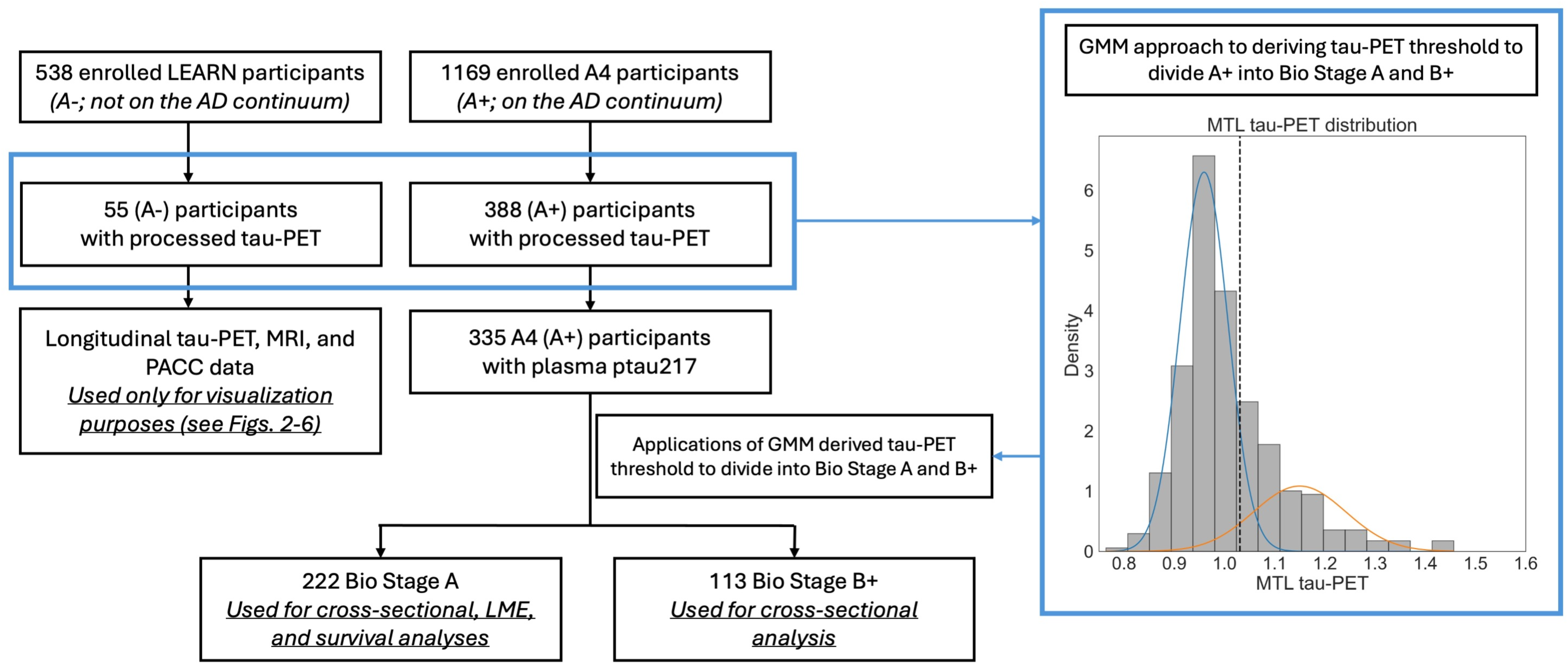
Flow chart illustrating selection of participants for analysis. From the A4 study, we included patients with complete amyloid-PET, tau-PET, and p-tau217 data. From the LEARN study, we included participants that had tau-PET data. Gaussian mixture modeling was applied to the tau-PET data from A4 and LEARN to derive an MTL tau-PET threshold. This threshold was applied to the A4 participants to derive Biological Stage A and Biological Stage B+ groups.

### Biomarker and Cognitive Measures

We used florbetapir-positron emission tomography (amyloid-PET), flortaucipir-PET (tau-PET), and T1-weighted magnetic resonance imaging (MRI) data. For amyloid-PET, a summary standard uptake value ratio (SUVr) was calculated for each participant using previously described methods^12,13^ (variable *Composite_Summary* from the spreadsheet *imaging_SUVR_amyloid.csv*), and converted to centiloids^14^. For tau-PET, regional tau burden was estimated by calculating regional target SUVrs from a template atlas derived from participants from the Harvard Aging Brain Study^15^ and normalized by the PERSI reference region^16^. For our statistical analyses (see below) we used regional tau data from entorhinal, parahippocampal, fusiform, and inferior temporal. These data were extracted from *imaging_SUVR_tau.csv*. For MRI, the T1-weighted images were processed locally using the FreeSurfer longitudinal processing stream^17^. For our analyses, we used regional thickness values from entorhinal, parahippocampal, fusiform, and inferior temporal.

We used baseline plasma p-tau217 levels (variable *ORRES*, spreadsheet *biomarker_p- tau217.csv*). Plasma p-tau217 levels were measured using an ECL immunoassay as previously described^18^. In the released data, some subjects had raw baseline plasma p-tau217 levels (variable *ORRESRAW*) that were below the limits of the assay’s quantification. These values were excluded for primary analyses, but included in a sensitivity analysis (see Supplemental Results 1).

To examine cognition, we used the preclinical Alzheimer cognitive composite (PACC) (variable *PACC*, spreadsheet *PACC.csv)*^19^.

### Identification of Biological Stage A Participants

Amyloid positivity for inclusion into A4 was determined using a hybrid quantitative/qualitative approach implemented by the A4 study team. Participants with a summary florbetapir SUVr >= 1.15 (corresponding to a centiloid value of 33.3) were considered amyloid positive (amyloid+); participants with summary SUVr 1.10-1.15 were also considered amyloid+ if two readers gave consensus positive reads. All participants enrolled in A4 were amyloid+ and thus considered to be on the AD continuum.

We divided A4 (amyloid+) participants into Biological Stage A and Stage B+ (Stage B or higher). Per the latest criteria, Stage B+ can be defined by amyloid+ in conjunction with MTL tau positivity detected with PET. To define MTL tau-PET positivity, we calculated a weighted average SUVr from bilateral entorhinal and parahippocampal cortex (variables *entorhinal_lh, entorhinal_rh, parahippocampal_lh, parahippocampal_rh*, spreadsheet *imaging_SUVR_tau*.csv) (Supplemental Methods 1). Gaussian mixture modeling (GMM) was applied to the MTL SUVr tau-PET distribution for all participants with tau-PET, using baseline MTL SUVr data only (Figure 1). The threshold was set at 1.5 standard deviations (SD) above the mean of the tau-negative component from the GMM (threshold=1.03). Importantly, we used a conservative 1.5 SD threshold (compared to, for example, 2 or 2.5 SD) to ensure removal of MTL tau-PET positive cases from Biological Stage A. Amyloid+ participants with an MTL SUVr below and above this threshold were Biological Stage A and B+, respectively.

### Statistical Analyses

We first performed cross-sectional correlational analyses to evaluate baseline pairwise associations between continuous centiloid, MTL tau-PET, and p-tau217. We performed these cross-sectional analyses within the entire amyloid+ sample (Biological Stage A and B+ combined) and then within the Biological Stage A subgroup.

Next, in our primary analyses, we ran a series of linear mixed effects (LME) models to examine whether baseline Core 1 biomarkers (plasma p-tau217 or centiloid) were associated with longitudinal change in downstream disease markers within Biological Stage A. Briefly, in each LME model, the outcome was either regional tau SUVr, regional thickness, or PACC score. For each model, the predictor variable of interest was p-tau217 and its interaction with time (Model 1a) or centiloid and its interaction with time (Model 1b). The models can be represented as:

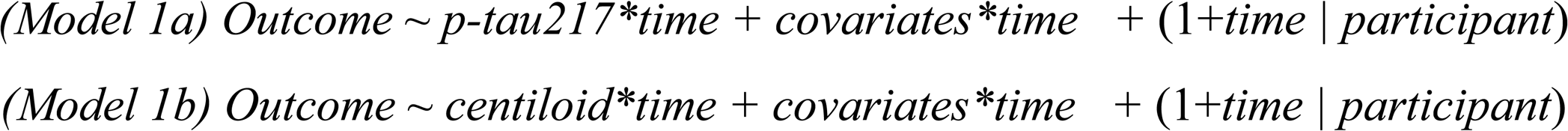

where *Outcome* is regional tau SUVrs (entorhinal, parahippocampal, fusiform, or inferior temporal), regional thickness (entorhinal, parahippocampal, fusiform, or inferior temporal), or PACC score. *Covariates* include age, sex, and APOE4 carriage. For the LME analysis in which PACC was the outcome, an additional covariate for education was also included. The (1+*time | participant*) term indicates that random slopes and intercepts were included in all models.

Then, we examined whether the Core 1 biomarkers were independently associated with longitudinal change in downstream markers:

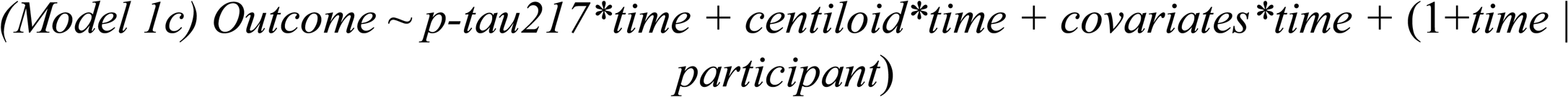

Estimates with p-value<0.05 were considered significant. The temporal lobe regions used in the LMEs were selected because of the predilection of tau and atrophy for these regions early in the disease course; further, they were assessed separately in the LMEs, rather than as a composite, to explore whether the effects of Core 1 markers on tau and atrophy may differ by regions involved very early (i.e., entorhinal and parahippocampal) versus early (i.e., fusiform and inferior temporal) in AD. To illustrate the findings from the LME analyses, we used a median split to divide participants into those with high and low p-tau217 or centiloid values, and then plotted the trajectories for these groups. Dichotomization of plasma p-tau217 and centiloid was done for visualization purposes only (all linear mixed models used continuous p-tau217 and centiloid levels). In sensitivity analyses, we also performed the LME analyses with covariates for race and drug group assignment (Supplemental Results 1).

Lastly, we used survival analyses to evaluate progression to Biological Stage B+ within our Biological Stage A sample. Cox proportional hazards modeling was used to assess the effect of continuous p-tau217 or centiloid values on the risk of progression to Biological Stage B+. P- tau217 values were z-scored so that hazard ratios can be interpreted as the risk associated with a 1 SD increase in p-tau217. Centiloid values were divided by 10 so that hazard ratios can be interpreted as risk associated with 10-point centiloid increase. An event was defined as a participant having a follow up tau-PET where their average MTL SUVr was above the threshold derived from the GMM procedure above. Following the same format as the LMEs (see above), We first evaluated p-tau217 and centiloid independently in separate models:

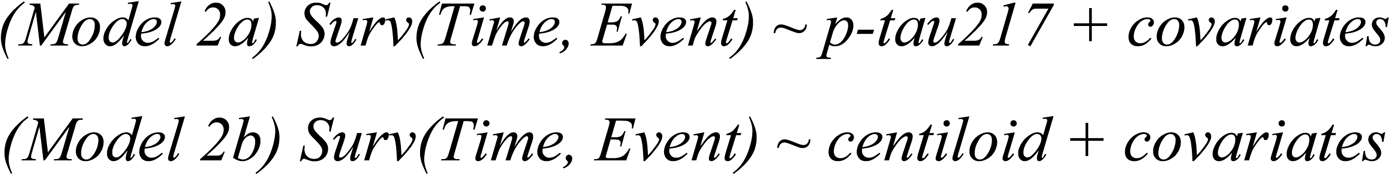

and then together in the same model to evaluate if they remained significant predictors of progression to Biological Stage B:

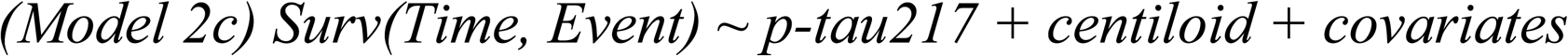

Lastly, Kaplan Meier plots were used to illustrate survival probabilities for participants based on p-tau217 or centiloid level. For Kaplan Meier plots, participants were divided into high and low p-tau217 groups as well as high and low centiloid groups using a median split.

## Results

### Participant Characteristics and Baseline Associations

There were 55 amyloid- (LEARN) participants with tau-PET data available and 335 amyloid+ (A4) participants with complete biomarker data (Figure 1). Among the amyloid+ group, 222 (66%) were classified as Biological Stage A and 113 (34%) as Biological Stage B+. Compared to Biological Stage A participants, Biological Stage B+ participants had higher continuous plasma p-tau217 (t=-6.55, p<0.001) and centiloids (t=-3.81, p<0.001). There were no statistically significant differences in age, sex, education, APOE genotype, race, or drug group assignment between Biological Stage A and B+ participant groups (Table 1).

**Table 1.** Subject demographics. Demographics for the A4 participants whose data were used in this study. Data are represented as mean (standard deviation) unless otherwise noted. T-tests were used to compare continuous variables across groups and Chi-squared tests were used to compare categorical variables across groups. Abbreviations: MTL=medial temporal lobe, PET=positron emission tomography, SUVr=standardized uptake value ratio, MRI=magnetic resonance imaging, PACC=preclinical Alzheimer’s cognitive composite.

|  | Biological Stage A | Biological Stage B+ |  |
| --- | --- | --- | --- |
| N | 222 | 113 |  |
| Age (years) | 71.9 (4.8) | 72.7 (4.8) | t=-1.43, p=0.16 |
| Sex (% female) | 55.0% | 61.9% | X=1.22, p=0.27 |
| Education (years) | 16.1 (3.0) | 16.4 (2.7) | t=-0.81, p=0.42 |
| APOE4 (% carriers) | 56.8% | 61.1% | X=0.41 p=0.52 |
| e2/e2 | 1 | 0 | X=5.35, p=0.37 |
| e2/e3 | 8 | 6 |  |
| e2/e4 | 8 | 2 |  |
| e3/e3 | 87 | 38 |  |
| e3/e4 | 106 | 55 |  |
| e4/e4 | 12 | 12 |  |
| Race |  |  |  |
| White | 205 | 99 | X=2.30 p=0.68 |
| Asian | 9 | 8 |  |
| Black | 5 | 3 |  |
| More than one race | 2 | 2 |  |
| Unknown | 2 | 1 |  |
| Treatment Group (% drug) | 47.7% | 54.0% | X=0.93 p=0.34 |
| MTL tau-PET (SUVr) | 0.96 (0.04) | 1.13 (0.09) | t=-23.5, p=1.38e-72 |
| Plasma p-tau217 | 0.25 (0.12) | 0.36 (0.18) | t=-6.55, p=2.15e-10 |
| Centiloid | 61.2 (28.4) | 74.7 (34.6) | t=-3.81, p=1.67e-4 |
| tau-PET follow up time (years) | 4.2 (2.1) | – |  |
| tau-PET scans (# of scans per participants) | 3.5 (1.3) | – |  |
| MRI follow up time (years) | 4.2 (1.9) | – |  |
| MRI scans (# of scans per participants) | 2.3 (0.77) | – |  |
| PACC follow up time | 5.7 (1.6) | – |  |
| PACC administrations | 11.6 (3.1) | – |  |

Significant cross-sectional associations were present between centiloid, plasma p-tau217, and MTL tau-PET, within the full amyloid+ sample (combined Biological Stage A and B+ combined), with shared variance ranging from 7 to 34% (p-tau217 vs. centiloid: R^2^=0.34, p<0.001; p-tau217 vs. MTL tau-PET: R^2^=0.19, p<0.001; centiloid vs. MTL tau-PET: R^2^=0.07, p<0.001)(Figure 2). Cross-sectional associations remained significant when restricting to the Biological Stage A group. Compared to the full amyloid+ sample, a similar amount of variance was shared between p-tau217 and centiloid (R^2^=0.31, p<0.001). However, the amount of variance shared between p-tau217 and MTL tau-PET (R^2^=0.04, p=0.006) as well as between centiloid and MTL tau-PET (R^2^=0.03, p=0.02) was reduced by more than 50% when restricted to Stage A.

**Figure 2.**
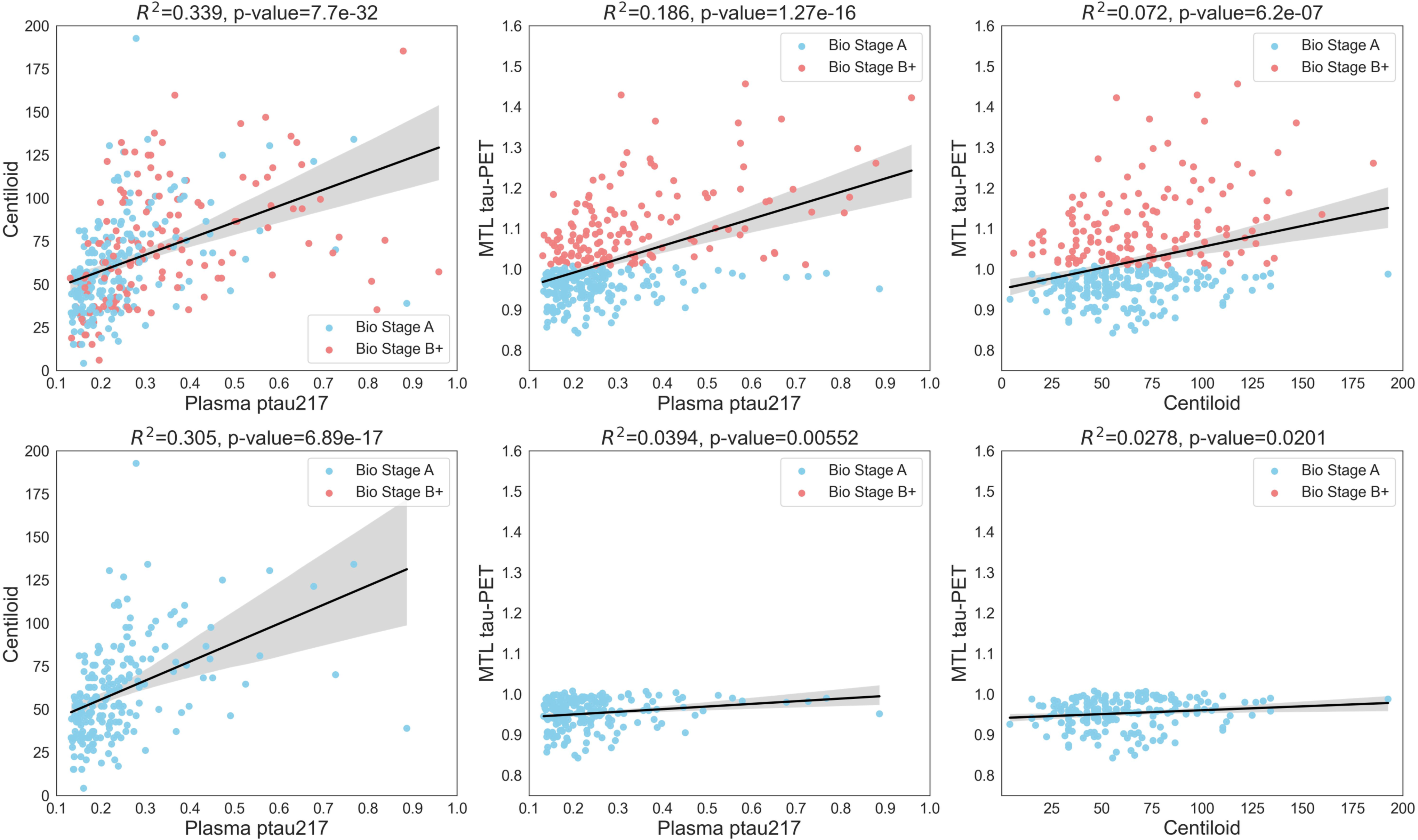
Cross sectional associations between plasma p-tau217, amyloid-PET, and tau-PET. Cross sectional centiloid, entorhinal tau, and p-tau217 were all significantly correlated with each other. In the top panel, both Bio Stage A and Bio Stage B+ participants are included in the pairwise analyses. In the bottom panel, only Bio Stage A participants are included in the analyses. R-squared and p-values are reported for each pairwise correlation.

### Prediction of tau accumulation, atrophy, and cognition the Biological Stage A

A series of LMEs were run within the Stage A group to determine whether baseline continuous plasma p-tau217 and centiloid values were associated with subsequent tau-PET accumulation, atrophy, and cognitive performance (Supplemental Table 1).

### Tau-PET accumulation

Baseline plasma p-tau217 (Model 1a) was associated with higher tau accumulation in all examined regions: entorhinal (β=0.0337, SE=0.009, p<0.001), parahippocampal (β=0.035, SE=0.008, p<0.001), fusiform (β=0.037, SE=0.01, p<0.001), and inferior temporal (β=0.049, SE=0.012, p<0.001) (Figure 3a). Likewise, baseline centiloid (Model 1b) was associated with higher tau accumulation in all regions of interest: entorhinal (β=8.61e-5, SE=3.84e-5, p=0.026), parahippocampal (β=1.19e-4, SE=3.07e-5, p<0.001), fusiform (β=1.51e- 4, SE=3.95e-5, p<0.001), and inferior temporal (β=2.38e-4, SE=4.61e-5, p<0.001) (Figure 3a).

**Figure 3.**
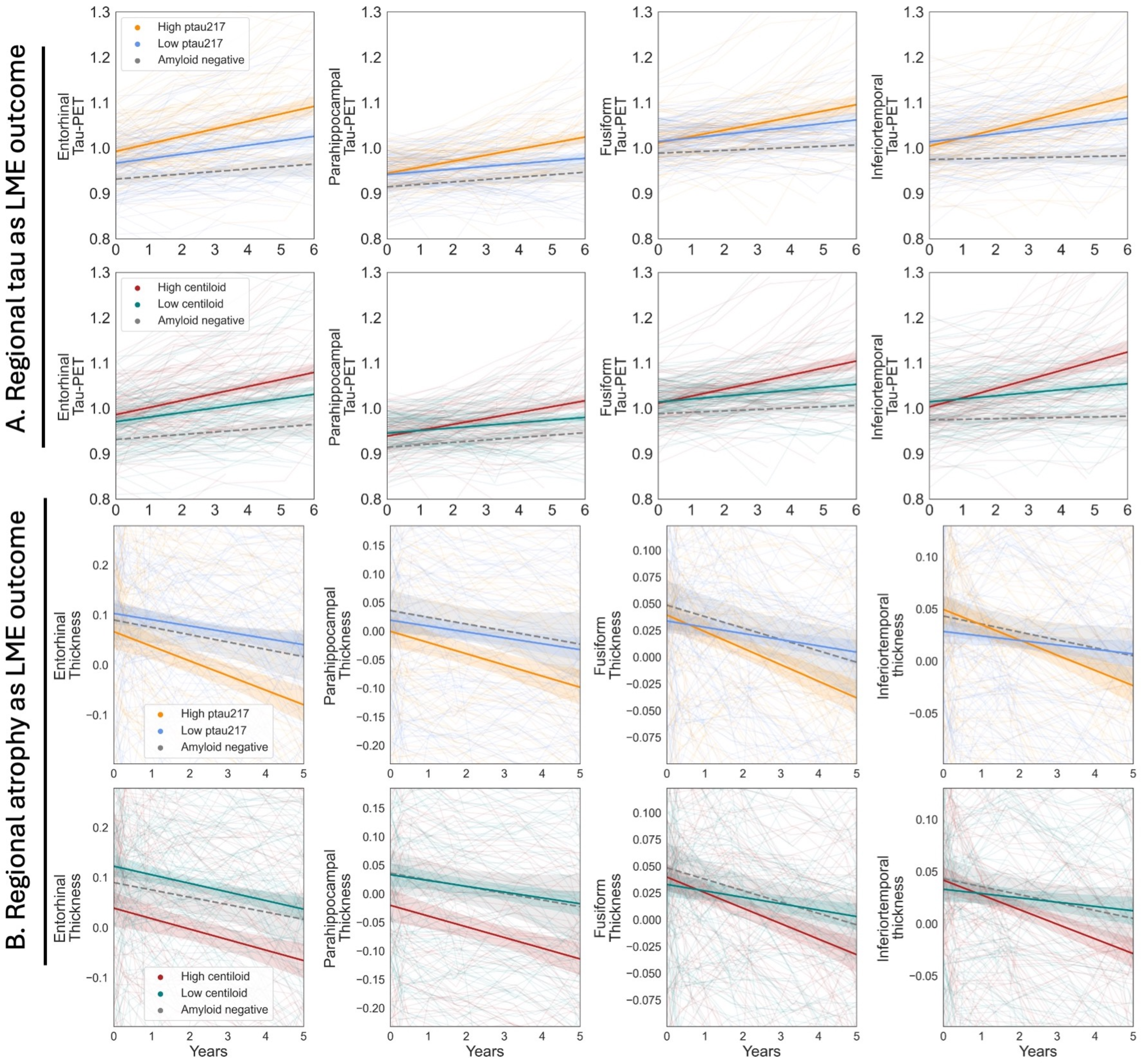
Higher p-tau217 and centiloid are associated with faster regional tau accumulation and worse atrophy. In panel (A), we show the relationship between plasma p-tau217 or centiloid with regional tau-PET. In panel (B), we show the relationship between plasma p-tau217 or centiloid with regional atrophy. In the plots, the orange and blue lines show the trajectories of high and low plasma p-tau217 participants, respectively.These groups were defined using a median split. The average p-tau217 level within the high and low p-tau217 subgroups is 0.17 and 0.33, respectively. The red and green lines show the trajectories of high and low centiloid participants, respectively. Again, a median split was used to define these groups. The average centiloid level within the high and low centiloid subgroups is 39.3 and 83.4, respectively. Binarized groups are shown here for visualization purposes, but in the LME analysis, continuous measures were used.

In LME models that included both baseline plasma p-tau217 and baseline centiloid as simultaneous predictors (Model 1c), plasma p-tau217 remained associated with regional tau accumulation (entorhinal, parahippocampal, and fusiform all p<0.05; inferior temporal trend level p=0.07). Baseline centiloid remained associated with tau accumulation in fusiform and inferior temporal (p<0.05), but not significantly associated with tau accumulation in entorhinal (p=0.73) and was trend level with parahippocampal (p=0.07).

### Atrophy

Baseline plasma p-tau217 (Model 1a) was associated with greater longitudinal atrophy across all regions: entorhinal (β=-0.064, SE=0.015, p<0.001), parahippocampal (β=-0.054, SE=0.009, p<0.001), fusiform (β=-0.039, SE=0.008, p<0.001), and inferior temporal (β=-0.052, SE=0.009, p<0.001) (Figure 3b). Likewise, baseline centiloid (Model 1b) was associated with greater atrophy in all regions: entorhinal (β=-1.4e-4, SE=6.32e-5, p=0.028), parahippocampal (β=-1.25e-4, SE=3.86e-5, p=0.001), fusiform (β=-1.26, SE=3.09e-5, p=6.42e-5), and inferior temporal (β=-1.78e-4, SE=3.55e-5, p=1.31e-6) (Figure 3b). In models that included both baseline plasma p-tau217 and centiloid as predictors (Model 1c), plasma p-tau217 remained significantly associated with greater atrophy in all four regions (p<0.05). Baseline centiloid remained associated with inferior temporal atrophy (p=0.01), but was no longer associated with entorhinal (p=0.83) or parahippocampal (p=0.62) atrophy. The association between centiloid and fusiform was trend level (p=0.07).

#### Cognitive performance

In separate models, baseline plasma p-tau217 (Model 1a; β=-1.65, SE=0.43, p<0.001) and baseline centiloid (Model 1b; β=-0.006, SE=0.002, p<0.001) were associated with worse longitudinal PACC performance (Figure 4). When modelled together (Model 1c) both baseline plasma p-tau217 (β=-1.16, SE=0.49, p=0.019) and centiloid (β=-0.004, SE=0.002, p=0.031) values remained significantly associated with worse longitudinal PACC scores.

**Figure 4.**
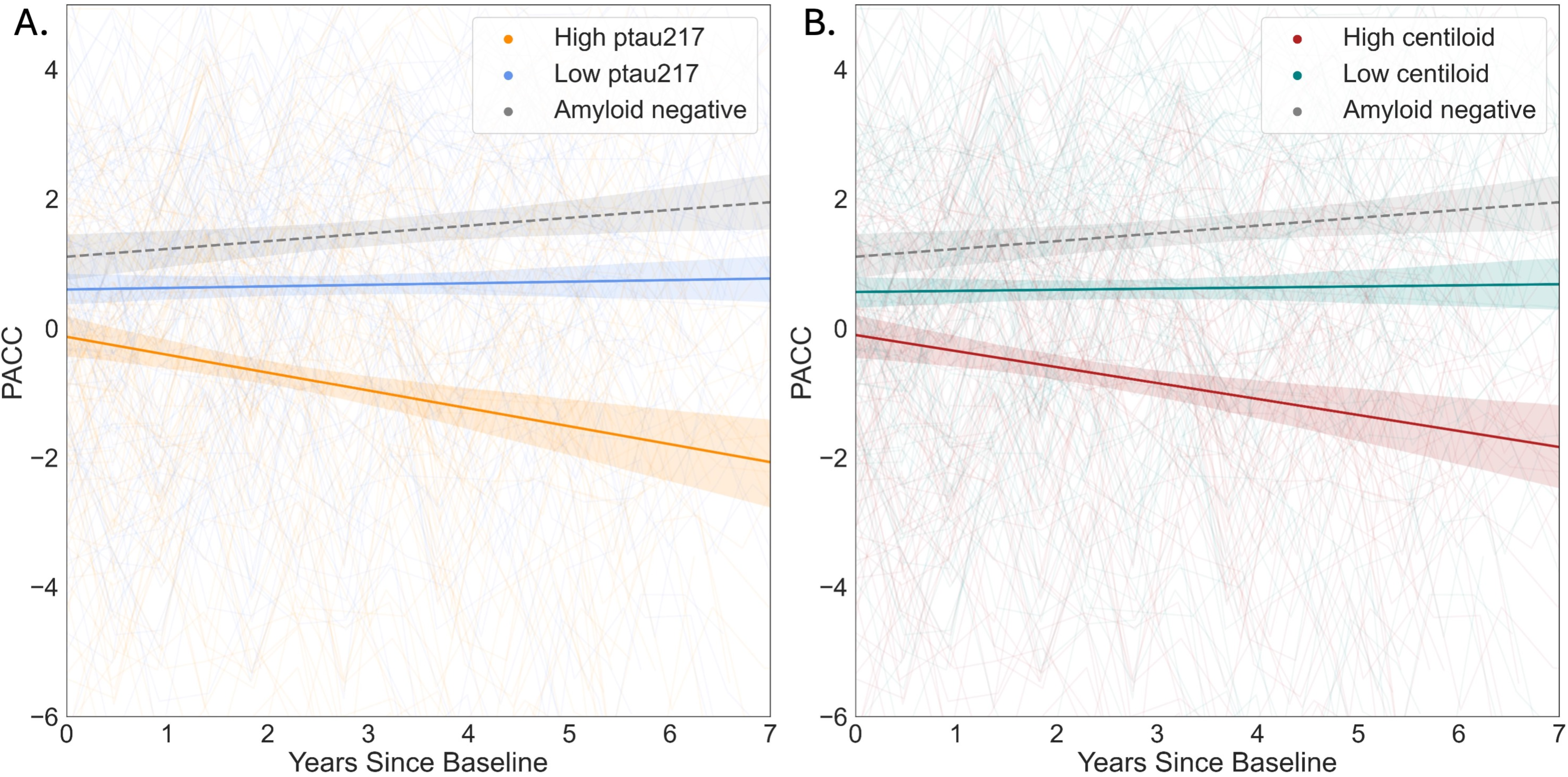
Higher p-tau217 and centiloid are associated with worse cognitive trajectory. In panel (A), we show the relationship between baseline plasma p-tau217 with longitudinal PACC. In panel (B), we show the relationship between baseline centiloid with longitudinal PACC. As was the case in Figure 3, the orange and blue lines show the trajectories of high and low plasma p-tau217 participants, respectively. The red and green lines show the trajectories of high and low centiloid participants, respectively. Again, these were defined by median splits. Binarized groups are shown here for visualization purposes, but in the LME analysis, continuous measures were used.

### Plasma p-tau217 and centiloid are associated with higher risk of progression to Biological Stage B+

A total of 48 Biological Stage A participants (22%) progressed to Biological Stage B+ during the course of follow-up (mean follow up=4.2 years; Figure 5). When examined in separate models, continuous p-tau217 (HR=1.81, CI=(1.41-2.32), p<0.001) and centiloid (HR=1.24, CI=(1.12- 1.38), p<0.001) were associated with increased progression to Biological Stage B+. Both p- tau217 and centiloid values were independently associated with progression to Biological Stage B+ when included in the same model (p-tau217: HR=1.49, CI=[1.10-2.03], p=0.01; centiloid: HR=1.15, CI=[1.03-1.30], p=0.02).

**Figure 5.**
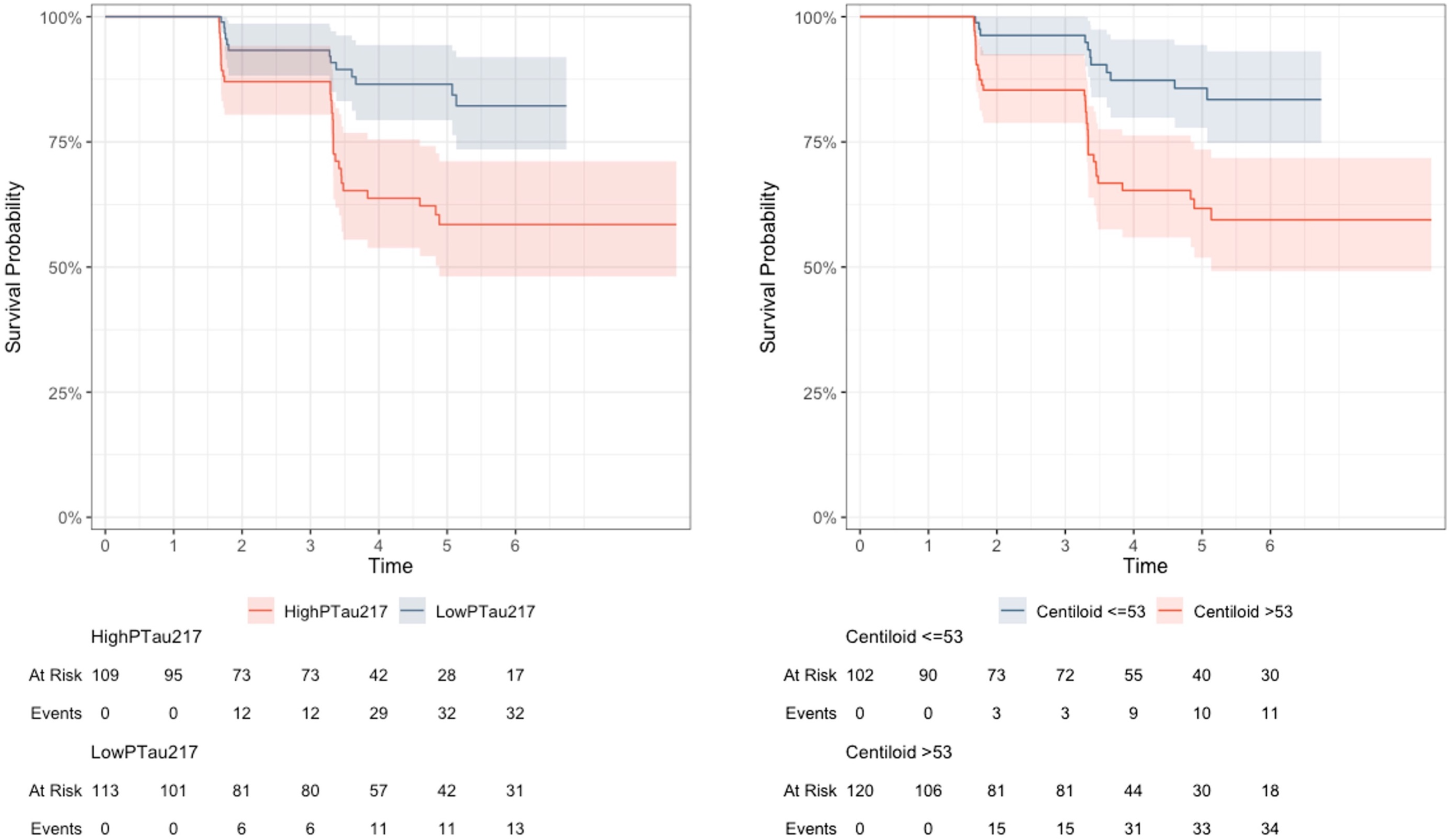
Higher p-tau217 is associated with higher risk of conversion to Biological Stage B. Graphs show the Kaplan-Meier estimates for survival based on p-tau217 (left panel) or centiloid (right panel). Definition for high p-tau217 is identical to that shown in Figure 3 and 4 (i.e., Biological Stage A participants with p-tau217 level greater than 2 SDs above the mean from the LEARN distribution). For centiloid, we used a median split to divide the sample into high and low centiloid groups. In the Cox proportional hazards analyses, continuous, rather than binary, measures of p-tau217 and centiloid were used.

## Discussion

In this study, we implemented the recently proposed 2024 criteria for the diagnosis and staging of AD to identify and characterize a group of CU individuals in Biological Stage A. This stage is reflective of the earliest biological stage of AD detectable with current biomarkers, reflecting a stage in which early pathological processes have been initiated. More specifically, this stage can be defined by positivity in “Core 1” early changing markers, such as amyloid-PET or plasma p- tau217, in conjunction with negativity in downstream tau markers that are indicative of disease progression. Within this early Stage A group, we sought to establish whether variability in continuous measures of early changing Core 1 markers were biologically relevant. We found that both baseline amyloid-PET and plasma p-tau217 were associated with longitudinal accumulation in tau-PET, greater atrophy, and worse cognitive performance. Complementing these longitudinal models, we also found that elevated baseline amyloid-PET and plasma p- tau217 were also independently associated with faster progression to Biological Stage B+ (MTL tau-PET positivity). Our findings highlight that variability within Core 1 AD biomarkers provides important prognostic information, even during the earliest stage of biologically defined AD.

Our finding that continuous levels of amyloid-PET and plasma p-tau217 are biologically meaningful is consistent with other studies. Within CU individuals, multiple studies have shown that continuous amyloid-PET signal (even within the A+ range) is associated with longitudinal cognitive performance or clinical progression to MCI or dementia ^4,20,21^. Similarly, multiple studies have also demonstrated that within cognitively unimpaired and A+ study participants, elevated levels of baseline plasma p-tau217 predicts longitudinal cognitive performance and clinical progression ^6,7^. Importantly, a recent A4 study showed that plasma p-tau217 was associated with longitudinal PACC performance among the amyloid+ CU ^5^. However, none of the aforementioned studies restricted analyses to the subgroup of amyloid+ CU individuals that were also negative on tau-PET, to establish whether the continuum of amyloid-PET and/or plasma p-tau217 remains biologically relevant. This is especially important to distinguish, as tau-PET+ CU are known to be at increased risk of future tau accumulation^22,23^ and clinical progression^24,25^. Our approach addresses this question, and additionally places findings in the context of the revised biological criteria for the diagnosis and staging of AD ^2^. Our results are the first to our knowledge to demonstrate that even among the earliest biologically defined stage of AD (Stage A), the continuum of both PET and plasma measures of early AD processes are predictive of multiple disease relevant changes over time. Notably, our finding that amyloid-PET and plasma p-tau217 independently predict decline in Biological Stage A is distinct from a prior study across the entire A4 cohort^5^ showing that when these two biomarkers were included in the same model, only p-tau217 contributed unique variance. It may be the case that in the earliest biological stage, both amyloid-PET and plasma p-tau217 carry unique prognostic value, but at a certain point in the disease process (e.g., Biological Stage B), the predictive effect of amyloid- PET plateaus and only plasma p-tau217 remains predictive of decline.

Plasma p-tau217 is among the newest of the staging biomarkers proposed for use in the 2024 criteria, and one assay measuring p-tau217 (different from the assay used in A4) has recently gained FDA clearance (as a ratio with plasma amyloid-β 42) ^26^. Early studies on plasma p-tau217 showed significant associations with established markers of amyloid and tau (i.e., CSF and PET)^27–30^. Further, multiple studies have demonstrated that plasma p-tau217 may be more strongly associated with amyloid than tau particularly early in the disease course ^27,28^, and therefore become abnormal earlier in the disease course ^31^. Given these early-changing characteristics, p-tau217 has been proposed to be a Core 1 biomarker in the revised criteria, to determine whether someone is on the AD spectrum (Stage A+) but not as a prognostic measure (Stages B-D). Plasma p-tau217 and amyloid-PET as continuous measures are strongly correlated, with AUCs typically exceeding 0.90 in discrimination analyses using continuous p-tau217 to predict amyloid-PET positivity ^30^. As a practical application, in the context of clinical trial screening, p-tau217 has been demonstrated to have promise for enriching preclinical AD trials for participants who are amyloid positive by PET imaging ^18,32–34^ . Overall, close coupling between continuous p-tau217 and amyloid-PET was recapitulated in our analyses even after restricting to the Stage A group. However, the shared variance between these two Core 1 measures was only around 30% among amyloid-PET+ individuals both with and without MTL tau positive cases in our analyses, and these measures likely reflect distinct aspects of early disease biology. Our main finding that both continuous centiloid and p-tau217 values were associated with multiple downstream markers highlights that each measure contains meaningful information, even within the earliest stage of AD.

While the associations of plasma p-tau217 and centiloids with downstream markers was relatively consistent across our analyses, there were some exceptions. Namely, in our LME analyses that included both centiloid and p-tau217 as simultaneous predictors, centiloids were not associated with longitudinal tau-PET or atrophy in the medial temporal regions whereas p- tau217 was a significant independent predictor. It is possible that the effects of baseline centiloid on downstream AD markers is mediated by plasma p-tau217, particularly in regions that are vulnerable very early in the course of AD, such as the medial temporal lobe. Data from other studies examining a broader range of participants beyond just Biological Stage A supports a model in which p-tau217 mediates the relationship between amyloid-PET and downstream markers such as tau-PET ^27,29,31,35^.

In revised criteria for the diagnosis and staging of AD, the workgroup explicitly states that the framework is meant to be applied only to those with cognitive impairment; further, they recommend that the staging be used for CU only in the context of research studies including clinical trials or observational studies. With a large focus in the field on secondary prevention (e.g., AHEAD 3-45 ^36^, TRAILBLAZER 3 [NCT05026866] , and SKYLINE [NCT05256134]), it is of critical importance to understand how this framework applies in the earliest biological stages in amyloid PET positive individuals without cognitive impairment. These findings are particularly relevant for trials aiming to move even earlier in the pathophysiological sequence, such as the AHEAD A3 trial at the intermediate levels of amyloid PET, and future trials getting closer to primary prevention of amyloid “positivity”. If a prevention trial is successful, medical practice will need to rapidly adapt and integrate biological staging into medical decision making for CU individuals. In addition to Core 1 biomarkers potentially being used in the future to confirm that an individual is on the AD neuropathological spectrum, our findings suggest that additional information leveraging the continuum of amyloid-PET and plasma p-tau217 values may be clinically relevant beyond just classification into Stage A. Future work in a real-world setting is critical to establish whether Core 1 biomarkers may be leveraged in a manner that provides individuals classified as Stage A with additional prognostic information.

Our study has multiple limitations. While this study is among the first to apply the new criteria to a large sample of CU older adults, the sample here is not representative of the general population given the high education in the cohort and the setting of a secondary prevention trial. Next, we defined Biological Stage A+ based on amyloid-PET, whereas plasma p-tau217 is also recommended for making this classification according to the new criteria. We focused on amyloid-PET positivity to define Stage A given that this classification was used for enrollment into A4 and involved a hybrid quantitative-qualitative method. Lastly, there exist AD variants in which tau pathology relatively spares the MTL and, to this point, prior work using the A4 dataset has identified participants with cortical, rather than MTL, predominant tau deposition ^37^. Since the division between Biological Stage A and B+ in our study was done based on MTL tau level (as is suggested by the newest staging criteria), this may have led to inclusion of cortical-predominant tau participants into Biological Stage A; plotting of distributions of tau level in cortical regions beyond the MTL demonstrates that none of our Biological Stage A participants had elevated tau-PET signal in the later Braak stages and thus it is unlikely that any tau cortical-predominant (i.e., MTL-sparing) participants were included in our Biological Stage A (Supplemental Results 2).

## Conclusion

Among CU participants in the earliest biological stage of AD, continuous levels of amyloid-PET and plasma p-tau217 independently predicted longitudinal change in downstream AD markers. This work highlights the potential for future strategies that leverage multiple early changing biomarkers of AD to inform individual level risk of future progression among CU.

## Supporting information

Supplemental Methods and Results

## Data Availability

All data produced in the present work are derived from public datasets that are available at https://www.a4studydata.org/ and https://www.synapse.org/a4_learn_datasharing/ after user registration.

## Acknowledgements

The A4 Study was a secondary prevention trial in preclinical Alzheimer’s disease, aiming to slow cognitive decline associated with brain amyloid accumulation in clinically normal older individuals. The A4 Study was funded by a public-private-philanthropic partnership, including funding from the National Institutes of Health-National Institute on Aging, Eli Lilly and Company, Alzheimer’s Association, Accelerating Medicines Partnership, GHR Foundation, an anonymous foundation, and additional private donors, with in-kind support from Avid Radiopharmaceuticals, Cogstate, Albert Einstein College of Medicine and the Foundation for Neurologic Diseases.The companion observational Longitudinal Evaluation of Amyloid Risk and Neurodegeneration (LEARN) Study was funded by the Alzheimer’s Association and GHR Foundation. The A4 and LEARN Studies were led by Dr. Reisa Sperling at Brigham and Women’s Hospital, Harvard Medical School, and Dr. Paul Aisen at the Alzheimer’s Therapeutic Research Institute (ATRI) at the University of Southern California. The A4 and LEARN Studies were coordinated by ATRI at the University of Southern California, and the data are made available under the auspices of Alzheimer’s Clinical Trial Consortium through the Global Research & Imaging Platform (GRIP). The complete A4 Study Team list is available on: https://www.actcinfo.org/a4-study-team-lists/. We would like to acknowledge the dedication of the study participants and their study partners who made the A4 and LEARN Studies possible.

