## Supplemental Methods and Results for "Continuum of Core 1 Biomarkers in Preclinical Alzheimer’s Disease"

Supplemental Methods 1. Calculation of tau-PET MTL SUVr.

Supplemental Methods 2. Derivation of time variable for linear mixed effects models analyses.

Supplemental Results 1. Sensitivity analyses using alternative biomarker thresholds and model covariates.

Supplemental Results 2. Average SUVr By Braak Region.

Supplemental Results 3. Cross sectional associations between plasma p-tau217 and inferior temporal or fusiform tau-PET.

**Supplemental Methods 1. Calculation of tau-PET MTL SUVr.**

To split the A4 sample into Biological Stage A and Biological Stage B+ (Stage B and beyond), we applied a Gaussian mixture model approach on the distribution of tau-PET MTL SUVrs. The tau-PET MTL SUVrs were defined as follows. For each participant, we extracted the regional SUVrs from bilateral entorhinal and parahippocampal cortex (variables *entorhinal_lh, entorhinal_rh, parahippocampal_lh, parahippocampal_rh*, spreadsheet *imaging_SUVR_tau*.csv) from their first tau-PET scan. We then calculated the MTL SUVr as the weighted average from these regional SUVrs. The weights are regional volumes derived from GTMseg, a FreeSurfer-based MNI-space atlas that was produced from averaging regional segmentations from the participants of the Harvard Aging Brain study. This was the same atlas from which the regional SUVrs above were derived. The atlas is available for download from <https://habs.mgh.harvard.edu/researchers/data-tools/>. Because this template atlas was used, the weights reflect atlas regional volumes, rather than A4 participant-level volumes.

**Supplemental Methods 2. Derivation of time variable for linear mixed effects models analyses.**

For tau-PET, MRI, and PACC data available on the A4 data portal, timing information is coded as a visit code *VISCODE* variable. This *VISCODE* variable had to be transformed into a time measurement for linear mixed effects models analyses. We linked the *VISCODE* variable with the days since consent *SVSDTC_DAYS_CONSENT* variable in the *SV.csv* spreadsheet. This linkage allowed us to calculate days between scans or PACC administrations. For tau-PET and MRI analyses, the *time* variable in the linear mixed effects models represents time since their first tau-PET or MRI scan. For PACC analyses, the *time* variable in the linear mixed effects models represents time since their baseline PACC administration.

There were 37 tau-PET scans that occurred at an “early termination” visit (*VISCODE* value 999) and had no corresponding days since consent *SVSDTC_DAYS_CONSENT* value. For these scans, we used the days since consent *SVSDTC_DAYS_CONSENT* value from *VISCODE* 997 (which corresponds to open label extension [early termination or study close]) as an estimate of time.

There were 5 tau-PET scans that occurred at other visits (*VISCODE* 27, 48, and 84) but had no days since consent *SVSDTC_DAYS_CONSENT.* To derive a time value for these scans, we took the *SVSDTC_DAYS_CONSENT* value from the prior *VISCODE* that had a corresponding *SVSDTC_DAYS_CONSENT* value. We then added the expected number of days based on the study schedule to get to the *VISCODE* of interest (see Table below for example calculation and for summary of these 5 cases; adapted from Young et al., 2025, *in preparation*).

| **BID** | **Missing *VISCODE*** | **Weeks at *VISCODE* (based on study schedule)** | **Prior *VISCODE* with corresponding *SVSTDTC_ DAYS_ CONSENT*** | **Weeks at prior *VISCODE* (based on study schedule)** | ***SVSTDTC_ DAYS_ CONSENT* at prior *VISCODE*** | **Estimated *SVSTDTC_ DAYS_ CONSENT*** |
| --- | --- | --- | --- | --- | --- | --- |
| B18613471 | 84 | 312 | 80 | 296 | 2353 | 2353 + (312 - 296)*7 = 2465 |
| B32880198 | 84 | 312 | 83 | 308 | 2358 | 3086 |
| B69161824 | 48 | 168 | 47 | 164 | 1310 | 1338 |
| B86248441 | 84 | 312 | 82 | 304 | 2239 | 2295 |
| B90395301 | 27 | 84 | 26 | 80 | 632 | 1248 |

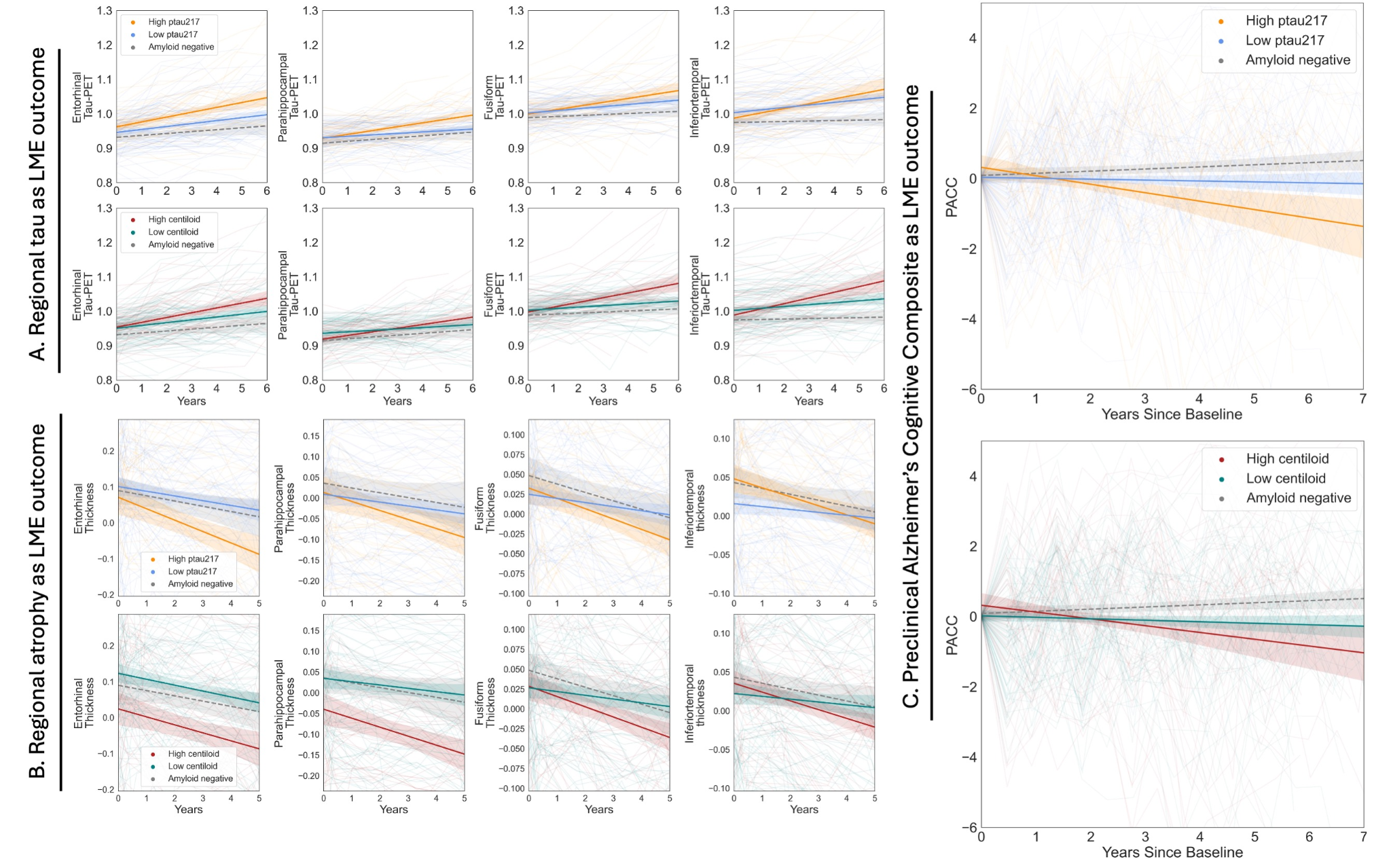

**Supplemental Results 1. Sensitivity analyses using alternative biomarker thresholds and model covariates.** In this sensitivity analysis, we used a threshold of 1 SD (rather than 1.5 SD) above the mean of the first component (tau-PET negative) identified by GMM to split the A4 sample into Biological Stage A and Biological Stage B+. We tested this more conservative threshold to ensure MTL tau-PET+ cases were excluded from Biological Stage A (i.e., to ensure our results were not being driven by a few subjects that had relatively higher MTL tau-PET+. In panel (A), we show the relationship between plasma p-tau217 or centiloid with regional tau-PET. In panel (B), we show the relationship between plasma p-tau217 or centiloid with regional atrophy. In panel (C) we show the relationship between plasma p-tau217 or centiloid. In the plots, the orange and blue lines show the trajectories of high and low plasma p-tau217 participants, respectively. These groups were defined using a median split. The red and green lines show the trajectories of high and low centiloid participants, respectively. Again, a median split was used to define these groups. Binarized groups are shown here for visualization purposes, but in the LME analysis, continuous measures were used.

Next, there are several participants that were included in A4 (deemed to have a positive amyloid-PET scan), but ultimately had a low calculated Centiloid level. In an additional sensitivity analysis, we excluded from our LME analyses participants that had a baseline Centiloid<=20.0. This excluded 8 participants and left 214 participants in Biological Stage A for analysis. The results largely remained unchanged.

Further, in our primary analyses, we excluded 23 participants with p-tau217 data below the limits of the assay, but with available raw data. Inclusion of the 23 participants with p-tau217 levels below the lower limit of the assay in our LME analyses produced similar results.

Lastly, we re-performed the LME analyses with different sets of covariates. The analyses were re-performed with a term to indicate treatment arm (Solanezumab or Placebo) and also re-performed without the APOE covariate term (since it may compete with amyloid-related variance in the linear mixed effects). The results largely remained unchanged.

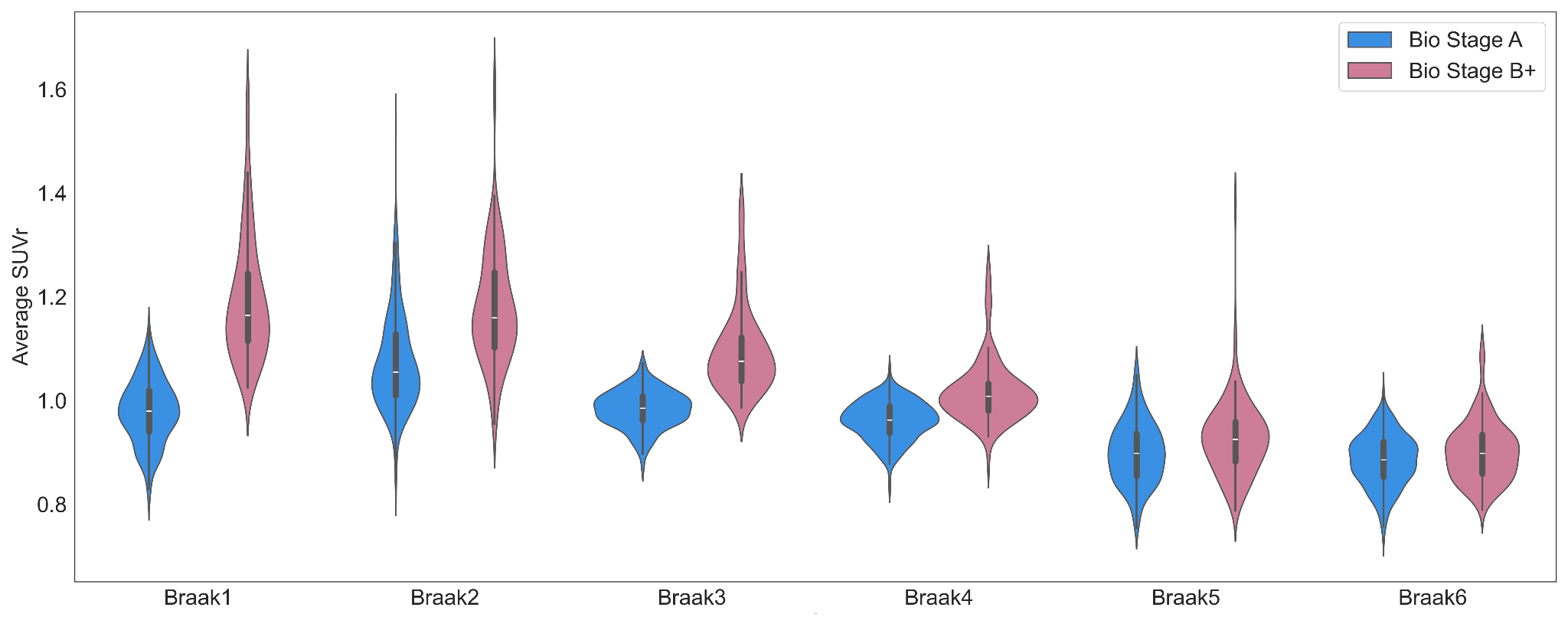

**Supplemental Results 2. Braak ROI SUVrs across Biological Stages.** Blue and red represent the distributions for the average Braak SUVr for the Biological Stage A and Biological Stage B+ groups, respectively. Each Braak SUVr is an average of the regional SUVrs of the regions that make up that specific Braak ROI (Scholl et al., 2016; PMID: 26938442). Given that the splitting of the sample into Bio Stage A and B+ was based on MTL tau-PET alone, we aimed to evaluate whether participants with hippocampal-sparing tau-PET patterns were included in the Biological Stage A group. Based on the distributions in the Braak 3-6 regions (which include neocortical regions), there is no indication that participants with hippocampal-sparing tau patterns were included into our Biological Stage A group. Abbreviations: ROI=region of interest; SUVr=standardized uptake value ratio; MTL=medial temporal lobe.

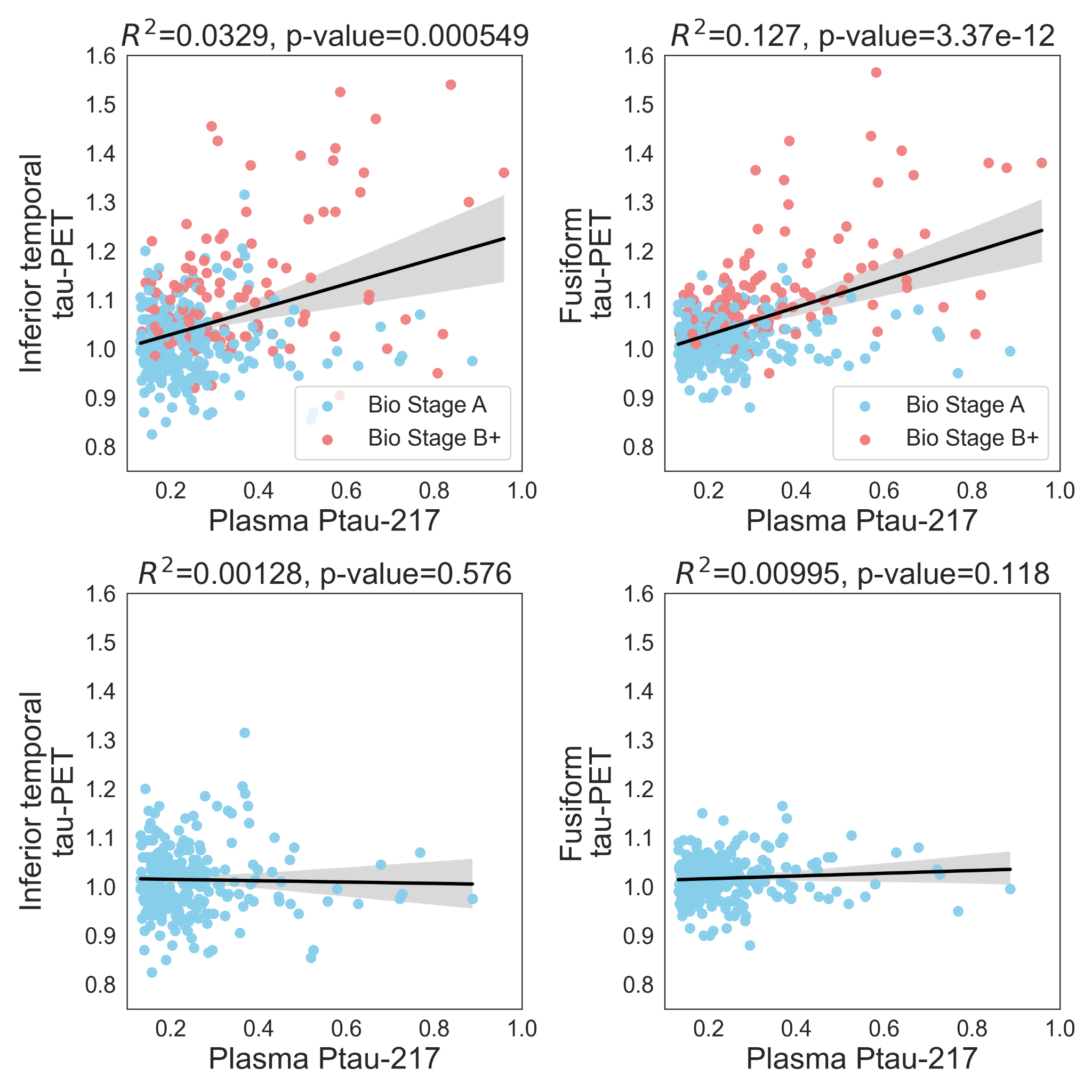

**Supplementary Results 3. Cross sectional associations between plasma p-tau217 and inferior temporal or fusiform tau-PET.** In the top panel, both Bio Stage A and Bio Stage B+ participants are included in the pairwise analyses. In the bottom panel, only Bio Stage A participants are included in the analyses. R-squared and p-values are reported for each pairwise correlation. This analysis was performed to visualize whether there were participants within our primary Bio Stage A group (amyloid-PET positive, MTL tau-PET negative group) that had very elevated tau-PET in temporal regions that are not part of our MTL meta-ROI (entorhinal and parahippocampal; we focused on entorhinal/parahippocampal since the new criteria focuses on medial temporal tau to define the boundary between Bio Stage A and B). Most of the Bio Stage A participants had inferior temporal and fusiform tau-PET values lower than the Bio Stage B+ participants, but there is some overlap between Bio Stage A and Bio Stage B+. Rather than applying GMM to MTL SUVrs (entorhinal / parahippocampal), we applied GMM to temporal meta-ROI SUVrs (entorhinal, parahippocampal, fusiform, inferior temporal, middle temporal, amygdala) to the boundary between Bio Stage A and B+. Defining the groups in this way, produced similar LME results to our primary LME analyses (*p-tau217*time* term p<0.05; data not shown). Abbreviations: MTL=Medial temporal lobe.
